# Chitinases in tear fluid of patients with Amyotrophic Lateral Sclerosis

**DOI:** 10.64898/2026.02.04.26345568

**Authors:** Lara Wenz, Lena-Sophie Scholl, Nya Reinhardt, Ricarda von Heynitz, Vincent Gmeiner, Petra Rau, Philipp Jie Müller, Emily Feneberg, Antonia F. Demleitner, Paul Lingor

**Affiliations:** Department of Neurology, Klinikum rechts der Isar, TUM Universitätsklinikum, School of Medicine and Health, Technical University of Munich, Munich, Germany; DZNE, German Center for Neurodegenerative Diseases, Munich, Germany; Munich Cluster for Systems Neurology (SyNergy), Munich Germany

**Keywords:** amyotrophic lateral sclerosis, biomarker, neuroinflammation, tear fluid, chitinase

## Abstract

**Background:** Chitinases, including chitotriosidase (CHIT1) and chitinase-3-like protein 1 (CHI3L1), are markers of neuroinflammation, a key process in amyotrophic lateral sclerosis (ALS). Tear fluid (TF) can be collected non-invasively and may represent a promising alternative to CSF or blood to study chitinases.

**Methods:** TF was collected from 50 ALS patients and 50 control subjects using Schirmer strips. CHIT1 and CHI3L1 levels in TF, serum, and CSF were quantified using ELISA. Serum NfL was measured using SIMOA. The frequency of a 24 bp-duplication polymorphism in the CHIT1 gene influencing CHIT1 expression was assessed by PCR.

**Results:** No group differences in the distribution of the CHIT1 polymorphism were detected. Carriers of the polymorphism in both ALS and controls showed lower CHIT1 levels in serum and TF. CHI3L1 levels in TF were higher in ALS patients compared to controls (p = 0.007), consistent with changes in CSF but not serum. In ALS, males showed higher TF CHIT1-values compared to females (p = 0.009). Combining TF chitinase values with serum NfL values improved discrimination between ALS and controls.

**Conclusions:** Chitinases are detectable in TF, and CHI3L1 levels recapitulate changes observed in CSF, highlighting its potential for non-invasive longitudinal assessment. Furthermore, chitinase values in TF, together with serum NfL, may act complementary by capturing distinct aspects of the disease, neuroinflammation and axonal damage. These results suggest TF chitinases and serum NfL could complementarily contribute to the diagnosis and monitoring of the disease, and call for further evaluation of TF as a biomarker source in ALS.

## INTRODUCTION

Amyotrophic Lateral Sclerosis (ALS) is a progressive neurodegenerative disorder characterized by the gradual degeneration and death of upper and lower motor neurons, which leads to muscle weakness and atrophy. It typically presents in mid to late adulthood, and the majority of patients succumb to the disease two to five years after symptom onset.^1^ The diagnosis of ALS is often delayed and remains challenging due to the considerable heterogeneity of the disease and unspecific initial symptoms.

Inflammatory processes, such as glial cell activation and infiltration of macrophages and T-lymphocytes, have been found to be upregulated in the disease course of ALS and are thought to play a key role in its pathogenesis.^2^ Clinical trials targeting inflammatory pathways have, however, not been successful in the general population of ALS patients. Recent studies on brain tissue and CSF suggest that inflammatory processes may be the dominant disease mechanism in only a subset of patients.^3-5^ This has further strengthened the view that ALS may represent not a single disease entity but rather a clinical syndrome comprising diverse clinical and molecular subtypes, one of which may be characterized by neuroinflammation.^6^ This underlines the urgent need for biomarkers identifying patients with prominent neuroinflammation for clinical interventions and to improve monitoring and outcome measurements of clinical trials.

Neurofilament light chain (NfL) is currently considered the most robust biomarker in ALS, offering reliable prognostic value and outperforming other candidates in distinguishing ALS from healthy controls, mimics, and other neurodegenerative diseases.^7^ Nonetheless, its lack of specificity, reflecting general axonal loss rather than specific pathophysiological processes, limits its utility in identifying distinct ALS subtypes. A potential biomarker candidate for the inflammatory subtype are chitinases, which have been shown to be associated with the activity of immune cells like macrophages and, in neurological contexts, microglial and possibly other glial cells^8^. Chitotriosidase (CHIT1) and chitinase-3-like protein 1 (CHI3L1) have been found to be increased in the cerebrospinal fluid (CSF) of patients with ALS across multiple studies (reviewed in ^9^). In addition, they have been linked to disease progression, disease severity, and patient survival (reviewed in ^10^). Results in serum are less conclusive, likely because chitinases are also abundantly expressed in peripheral, non-brain regions. Furthermore, serum is much more affected by systemic inflammatory processes like asthma and sarcoidosis, thereby reducing its specificity for CNS-related disease mechanisms.^11, 12^ CHIT1 levels in CSF as well as in serum are also influenced by a common 24-base pair duplication polymorphism in the CHIT1 gene, which leads to lower concentrations in heterozygous and often undetectable levels in homozygous carriers.^13, 14^

Collecting CSF is an invasive procedure and can be associated with relevant side effects and thus limits longitudinal assessments. Alternatively, less invasive biofluids like tear fluid (TF) have gained increasing attention as an accessible source of biomarkers for various diseases.^15^ Although TF originates as an ultrafiltrate from blood, the lacrimal gland, via parasympathetic innervation, also receives input from regions in the brain stem, which are areas commonly affected in many neurodegenerative disorders. Reduced TF production has been observed across several neurodegenerative diseases, potentially indicating an influence of neurodegeneration in the aforementioned regions on TF production.^16^ Furthermore, recent studies have reported changes in expression patterns of established biomarkers in TF of patients with Parkinson’s disease, Creutzfeldt-Jakob disease, and Huntington’s disease.^17-20^ Recently, analyses of the TF proteome in ALS patients and healthy controls revealed significant differences in expression using a biomarker signature comprising haptoglobin, serpin C1, and four other proteins.^21^

This study aimed to assess CHIT1 and CHI3L1 levels in the TF of patients with ALS and healthy control patients. We correlate the values with those in CSF and serum, as well as clinical and demographic parameters. For assessment of diagnostic performance, we further measured the corresponding serum NfL values and explored the potential additional diagnostic power of chitinases in TF and the other biofluids.

## MATERIALS AND METHODS

### Study design and participants

For this monocentric study, patients were recruited between September 2019 and January 2025 at the TUM University Hospital. Samples were collected from 50 patients with ALS who met the Gold Coast criteria^22^ at the time of sampling and 50 control subjects. The control group consisted of patients without evidence of neurodegenerative diseases. No other inclusion or exclusion criteria regarding age, sex, disease duration, concomitant diseases, or systemic or topical medication were applied. The disease progression rate for ALS patients was calculated as 48 minus the ALS-FRS-R at the time of sampling, divided by the disease duration in months. The characteristics of both groups are shown in Table 1 and Supplementary Table 1 in detail.

Written informed consent was obtained from all participants. The study complies with the Declaration of Helsinki and was approved by the Ethics Committee of the Technical University of Munich, School of Medicine (approval numbers: 9/15S, 2021-473-S-KH).

### Sample collection and processing

TF samples were collected according to a previously published protocol^16^ using an unanesthetized Schirmer strips (Madhu Instruments Pvt. Ltd., New Delhi, India) (for details see Supplementary Methods). Wetting length (WL) as well as clinical data (ophthalmological diseases, eye medication, and the use of contact lenses) were noted. Protein isolation was carried out as previously published. ^21^

CSF and serum were collected for diagnostic purposes and treated according to routine protocols.

### PCR genotyping of the CHIT1 24 bp duplication polymorphism

The patients’ DNA was extracted from 3 mL of blood using standard procedures. To detect the CHIT1 24 bp duplication polymorphism, PCR was performed using the following, previously published, primers: CHIT_ex10 for 5’-AGC TAT CTG AAG CAG AAG and CHIT_ex10rev 5’-GGA GAA GCC GGC AAA GTC, followed by electrophoresis on a 4% agarose gel. ^23^ The genotype was determined based on the length of the DNA fragments (75 and/or 99bp).

### Biochemical assays

Chitinase levels in all three biofluids were quantified using commercially available ELISA kits. Kits were tested for efficiency for use with TF. For CHIT1, the CircuLex human chitotriosidase ELISA (MBL, UK) was used according to the manufacturer’s instructions. Samples were measured in duplicates with an internal control and diluted 1:2 for TF, 1:50 for serum, and 1:10 for CSF samples. Likewise, for CHI3L1, the human chitinase-3-like 1 Quantikine ELISA kit (R&D Systems, UK) was used, and samples were diluted 1:4, 1:50, and 1:200 for TF, serum, and CSF, respectively. For samples with values below the lower limit of quantification (LLOQ) but above the limit of detection (LOD), the value was set at half of the value of the LLOQ, whereas for samples with values below the LOD, the value was set to zero. One control patient showed abnormally high CHI3L1-values in serum (1325.83 ng/ml), almost 30 times higher than the average concentration, and was therefore excluded from all further analyses after testing for outliers using Grubb’s test.

Total protein concentration in TF was determined by BCA to adjust the measured protein levels in TF accordingly.

NfL-values in serum were measured according to the manufacturer’s instructions using the Simoa® NF-light™ Advantage Kit Lot number 503196 (Quanterix, USA).

### Statistical and data analysis

All statistical analysis was performed using R version 4.4.1 (The R Foundation for Statistical Computing, Vienna, Austria). A p-value of below 0.05 (5%) was considered statistically significant. Categorical variables were compared using Pearson’s chi-squared test. For comparisons of continuous variables between two groups, the p-value was determined by Wilcoxon test, whereas for comparisons with more than two groups Kruskal-Wallis test with Dunn’s post-hoc and Holm correction was used. Prior, all groups were tested for normality by Shapiro-Wilk-test. For comparisons of TF wetting length (WL) between the groups, the cumulative WL of both eyes (in mm/5 min) was calculated, and a multiple linear regression correcting for the confounding variables age and sex was performed. Linear correlations between variables were assessed using Pearson’s correlation analysis. Classifier performance was evaluated by receiver operating characteristic (ROC) with the corresponding value of the area under the curve (AUC) and confidence intervals (CI), calculated according to the DeLong-Method. For visualization scales were pseudo-logarithmized.

## RESULTS

### Descriptive analysis of the study cohort

TF from a total of 100 patients, of which 50 were ALS patients and 50 controls, were collected and analyzed. Table 1 shows all demographic data as well as biomarker abundance values in detail. Both groups were well matched in regard to age, sex, ocular disease, and medication. Importantly for the CSF analyses, CSF standard parameters such as cell count, glucose, lactate, and albumin ratio were comparable between groups (Supplementary Table 1). Tear fluid production quantified by wetting length was reduced in the ALS group compared to the control group using a multiple linear regression model correcting for age and sex (ALS mean = 17.38 ± 17.20 mm/5 min, control mean = 25.60 ± 18.01 mm/5 min, estimate = -8.56, 95% CI [-15.47, -1.64], p = 0.016). The wetting length from all patients correlated strongly with the total protein concentration in TF (R_P_ = 0.75, 95% CI [0.64, 0.82], p < 0.001, Supplementary Figure 1).

### Analysis of the CHIT1-24bp-polymorphism

We assessed the presence of a 24bp duplication polymorphism, which has been reported to cause lower expression levels of CHIT1 in heterozygous and homozygous carriers (10.1007/s00415-024-12699-1, 10.1016/j.cca.2020.11.025). The control group consisted of 34 wild-types (72%) and 13 heterozygotes (28%), whereas the ALS group consisted of 33 wild-types (66%), 13 heterozygotes (26%), and 4 homozygotes (8%). This distribution shows a tendency towards a higher occurrence of the mutation in the ALS group (Figure 1a).

**Figure 1:**
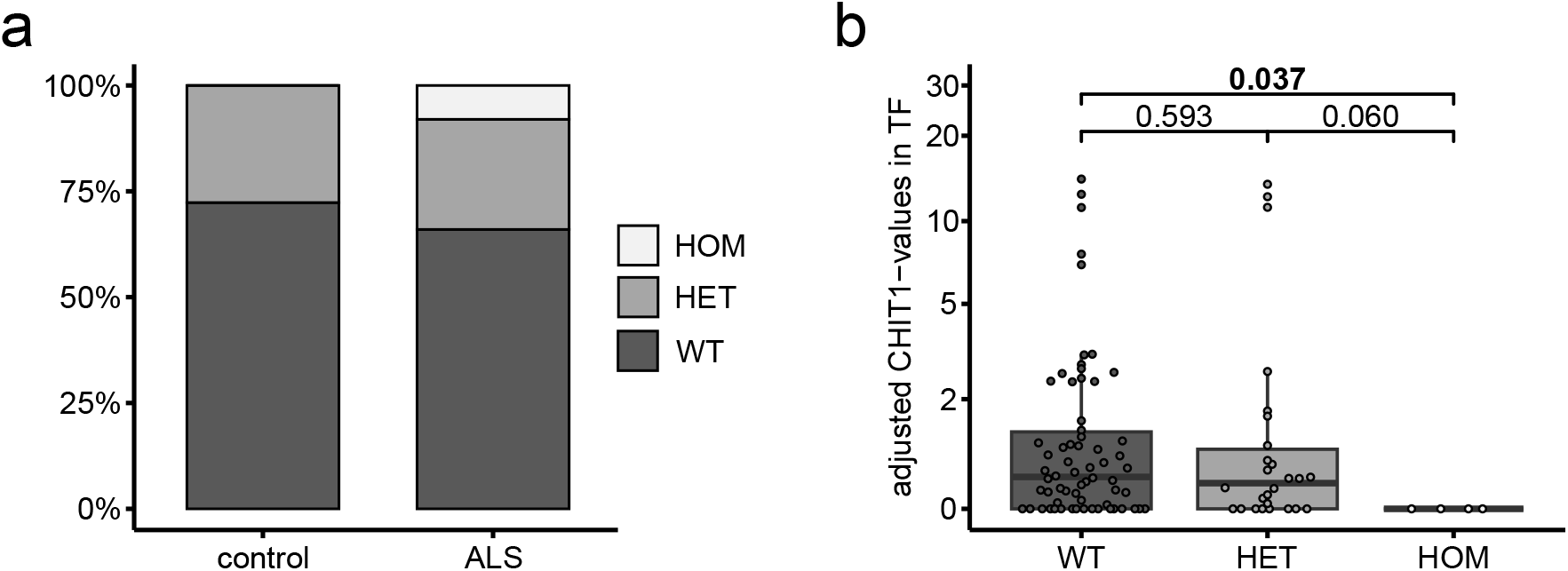
Analysis of the CHIT1 24bp duplication polymorphism. **a** Distribution of the CHIT1 polymorphism (wild-type, heterozygous, and homozygous) in the ALS (*n* = 47) and control group (*n* = 50) in percent. **b** CHIT1-values in TF depending on the CHIT1-polymorphism (wild-type, *n* = 66; heterozygous, *n* = 36; homozygous, *n* = 4). Subgroups were compared by Kruskal-Wallis test followed by Dunn’s post-hoc test adjusted for multiple comparisons with Holm correction. Absolute p-values are depicted. Boxplots for all graphs show median and 1st and 3rd quartile as well as all individual data points. TF-values were adjusted for total protein concentration. Abbreviations: CHIT1, chitotriosidase; CHI3L1, chitinase-3-like protein 1; ALS, amyotrophic lateral sclerosis; TF, tear fluid; WT, wild-type; HET, heterozygous; HOM, homozygous.

In the serum of carriers of the polymorphism, both heterozygotes and homozygotes showed significantly lower CHIT1 expression levels compared to wild-types (heterozygotes mean = 47.03 ± 69.69 ng/ml, homozygotes mean = 6.81 ± 13.63 ng/ml, wild-type mean = 56.64 ± 46.52 ng/ml, p_WTvsHET_ = 0.006, p_WTvsHOM_ = 0.001, Supplementary Figure 2). The expression levels between heterozygotes and homozygotes also showed a tendency towards lower levels in the latter, close to significance (p_HETvsHOM_ = 0.062).

Similarly, in TF, wildtype carriers showed the highest expression of CHIT1, and differed significantly from the levels of homozygous carriers, although not from heterozygous carriers (wildtype mean = 1.53 ± 2.84, homozygotes mean = 0.00 ± 0.00, heterozygotes mean = 1.88 ± 3.91, p_WTvsHOM_ = 0.037). Like in serum, heterozygous carriers also showed a trend towards higher levels of CHIT1 compared to homozygous carriers, though this difference did not reach significance (Figure 1b).

### Expression levels of CHIT1 and CHI3L1 in all three biofluids

The concentrations of the two chitinases, CHIT1 and CHI3L1, were determined in all three biofluids (TF *n* = 100, serum *n* = 100, CSF *n* = 28). TF values of CHIT1 or CHI3L1 did not significantly correlate with the corresponding values in serum or CSF (Supplementary Figure 3a-d). Likewise, no significant correlation was observed between serum and CSF values (Supplementary Figure 3e, f).

TF concentration of CHIT1 did not differ significantly between the ALS and control group, although concentrations were numerically higher in the ALS group (ALS mean = 1.87 ± 3.51, control = 1.23 ± 2.53). In CSF, CHIT1 levels of the ALS group were significantly higher than those of the control group (ALS mean = 15.50 ± 10.79 ng/ml, control mean = 6.47 ± 12.79 ng/ml, p < 0.001), while in serum no difference was found (ALS mean = 56.90 ± 62.92 ng/ml, control = 48.06 ± 41.30 ng/ml, Figure 2a-c).

**Figure 2:**
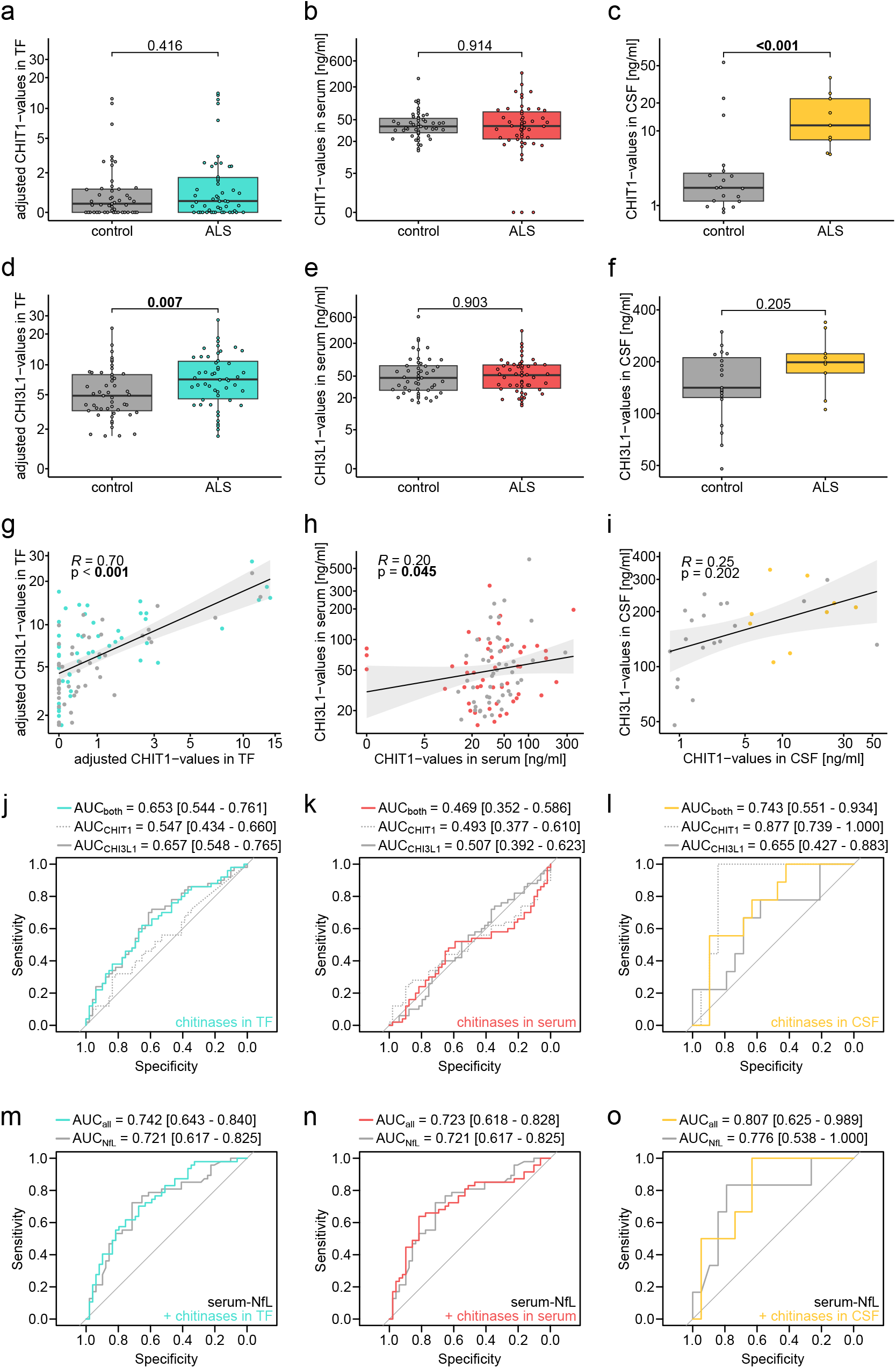
Comparison of chitinase levels in ALS-patients and controls in TF, serum and CSF. **a, b, c** CHIT1-levels of ALS-patients (a = turquoise, b = red, c = yellow) and controls (grey) in TF (**a**; ALS *n* = 50, control *n* = 49), serum (**b**; ALS *n* = 50, control *n* = 49) and CSF (**c**; ALS *n* = 9, control *n* = 19). Boxplots for all graphs show median and 1st and 3rd quartiles as well as all individual data points. **d, e, f** CHI3L1-levels of ALS-patients (d = turquoise, e = red, f = yellow) and controls (grey) in TF (**d**), serum (**e**) and CSF (**f**). **g, h, i** Pearson’s Correlation of CHIT1- and CHI3L1-values in TF (**g**), serum (**h**) and CSF (**i**). The regression line is shown in black with 95% confidence interval as well as all individual data points. Pearson’s correlation coefficient was calculated and is depicted at the top with corresponding p-values. **j, k, l** ROC-analysis (ALS vs. controls) of the CHIT1-(dotted grey line) and CHI3L1-values (continuous grey line) separately as well as both together in TF (**j**, turquoise line), serum (**k**, red line) and CSF (**l**, yellow line). **m, n, o** ROC-analysis (ALS vs. controls) of NfL-values in serum (grey line) as well as in combination with both chitinase-values in TF (**m**, turquoise line), serum (**n**, red line) and CSF (**o**, yellow line). AUC values with the 95% confidence interval are depicted. Groups were compared by Wilcoxon test/Mann-Whitney-U test. Absolute p-values and AUC values are depicted. TF-values were adjusted for total protein concentration before all analyses. Abbreviations: CHIT1, chitotriosidase; CHI3L1, chitinase-3-like protein 1; NfL, neurofilament light chain; ALS, amyotrophic lateral sclerosis; TF, tear fluid; CSF, cerebrospinal fluid; ROC, receiver operating characteristic; AUC, area under the curve.

CHI3L1 in the TF of ALS patients was significantly higher compared to healthy controls (ALS mean = 8.22 ± 5.01, control = 5.96 ± 4.06, p = 0.007). Similarly, in CSF ALS patients showed numerically, yet not significantly higher values in CHI3L1 (ALS mean = 208.48 ± 78.15 ng/ml, control = 159.27 ± 67.16 ng/ml). In serum, no difference was detectable between the ALS and control group (ALS mean = 64.17 ± 55.71 ng/ml, control mean = 73.88 ± 94.28 ng/ml, Figure 2d-f).

The correlation of CHIT1- and CHI3L1-values in TF was strong and significant, whereas it was weak and significant in serum. No significant correlation was found in CSF, although the number of available samples was markedly lower in the CSF group (TF: R_P_ = 0.70, 95% CI [0.59, 0.79], p < 0.001, serum: R_P_ = 0.20, 95% CI [0.00, 0.38], p = 0.045, Figure 2g-i, Supplementary Table 1). Neither of the chitinases in TF correlated with any of the clinical parameters, namely the ALS-FRS-R score, vital capacity, disease duration, or progression rate (Supplementary Figure 4).

Employing ROC-curve analysis, CHI3L1 in TF reached an area under the curve (AUC) of 0.657 for the discrimination of ALS from control patients, whilst CHIT1 achieved an AUC of 0.547 (CHI3L1: 95% CI [0.548, 0.765], CHIT1: 95% CI [0.434, 0.660]). The AUC of both chitinases together in TF did not outperform that of only CHI3L1 (AUC = 0.653, 95% CI [0.544, 0.761], Figure 2j). Neither the AUC-values of the chitinases separately nor together in serum outperformed the values in TF (CHI3L1: AUC = 0.507, 95% CI [0.392, 0.623], CHIT1: AUC = 0.493, 95% CI [0.377, 0.610], both: AUC = 0.469, 95% CI [0.352, 0.586], Figure 2k). CSF outperformed both other biofluids with CHIT1 as well as both chitinases together, whereas for CHI3L1 the AUC-value was below that of TF (CHIT1: 0.877, 95% CI [0.739, 1], both: AUC = 0.743, 95% CI [0.551, 0.934], CHI3L1: AUC = 0.655, 95% CI [0.427, 0.883], Figure 2l).

### Association between chitinase expression values and serum-NfL values

NfL was measured in the serum of all but three ALS patients. Neither of the chitinases in TF nor serum correlated with the NfL values in serum. In CSF, however, both chitinases showed a strong and significant correlation (CHIT1: R_P_ = 0.64, 95% CI [0.33, 0.83], p < 0.001, CHI3L1: R_P_ = 0.57, 95% CI [0.23, 0.79], p = 0.003, Supplementary Figure 5).

NfL in serum reached an AUC of 0.721 to distinguish ALS patients from controls (95% CI [0.617, 0.825]). In combination with the expression values of both chitinases in TF the performance was marginally improved to 0.742 (95% CI [0.643, 0.840]), whereas with the chitinase values in serum, it remained largely unchanged (AUC = 0.723, 95% CI [0.618, 0.828], Figure 2m, n). Considering only the patients with available CSF data (n = 28), serum NfL performed better (AUC = 0.776, 95% CI [0.538 -1.000]). Similarly, the combination with values of both chitinases in CSF showed an improvement in AUC (AUC = 0.807, 95% CI [0.625 -0.989], Figure 2o).

### Sex differences in TF

The CHIT1 values in the TF of the male subgroup in the whole cohort as well as in the ALS subgroup were significantly higher compared to the female subgroup (overall male mean = 2.38 ± 3.91, overall female = 0.56 ± 0.80, p = 0.009; ALS male = 3.26 ± 4.55, ALS female = 0.48 ± 0.62, p = 0.027; control male = 1.63 ± 3.15, control female = 0.66 ± 0.99, Figure 3a). This difference was not significant for CHI3L1-values. Here, the male subgroup showed a numerically higher expression levels between ALS and control patients compared to the female subgroup (overall male mean = 7.65 ± 5.38, overall female = 6.45 ± 3.63, ALS male = 9.35 ± 5.83, ALS female = 7.09 ± 3.84, control male = 6.17 ± 4.57, control female = 5.65 ± 3.28, Figure 3b). In the ALS subgroup, CHIT1-values also correlated significantly with the progression rate of ALS in males, whereas the CHIT1-values in females and neither of the CHI3L1-subgroups showed any correlation (R_P_ = 0.41, 95% CI [0.01, 0.69], p = 0.044, Figure 3c, d). Moreover, CHIT1 and CH3L1 separately as well as together performed better in distinguishing between ALS patients and healthy controls in the male subgroup compared to the female (CHIT1: AUC_male_ = 0.632, 95% CI [0.480, 0.783], AUC_female_ = 0.514, 95% CI [0.344, 0.684]; CHI3L1: AUC_male_ = 0.697, 95% CI [0.555, 0.838], AUC_female_ = 0.606, 95% CI [0.435, 0.777]; both: AUC_male_ = 0.690, 95% CI [0.547, 0.837], AUC_female_ = 0.656, 95% CI [0.491, 0.821]; Supplementary Figure 6).

**Figure 3:**
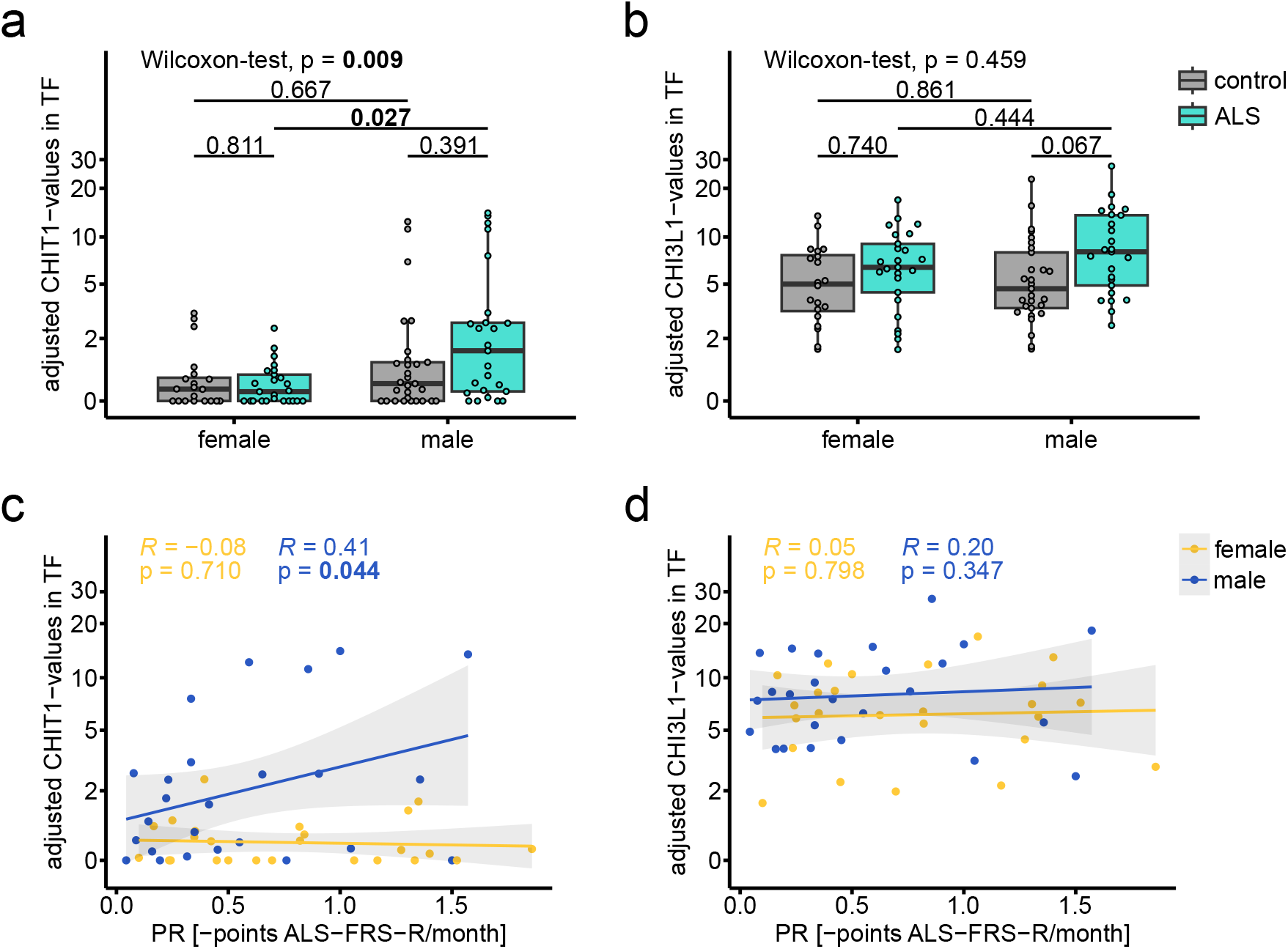
Sex differences in chitinase levels and correlations progression rate. **a, b** CHIT1-(**a**) and CHI3L1-values (**b**) in TF in males and females, further separated in the ALS (turquoise; female *n* = 25, male *n* = 25) and control (grey; female *n* = 20, male *n* = 29) group. Subgroups were compared by Kruskal-Wallis test followed by Dunn’s post-hoc test adjusted for multiple comparisons with the Holm correction. Absolute p-values are depicted (overall sex comparison by Wilcoxon test at the very top, subgroup comparisons by Kruskal-Wallis test with Dunn’s post-hoc and Holm correction below). Boxplots for all graphs show median and 1st and 3rd quartiles as well as all individual data points. **c, d** Pearson’s correlation of the progression rate of ALS-patients and CHIT1-(**c**) and CHI3L1-values (**d**) in TF stratified by sex. The regression line is shown in blue (males) and yellow (females) with a 95% confidence interval as well as all individual data points. Pearson’s correlation coefficient was calculated and is depicted at the top with corresponding p-values. TF-values were adjusted for total protein concentration. Abbreviations: CHIT1, chitotriosidase; CHI3L1, chitinase-3-like protein 1; ALS, amyotrophic lateral sclerosis; TF, tear fluid; ALS-FRS-R, revised amyotrophic lateral sclerosis functional rating scale.

## DISCUSSION

This study investigated the abundance levels of CHIT1 and CHI3L1 in the TF of ALS and control patients, and compared their levels to those in serum, CSF, and additional clinical parameters. As previously described, patients with ALS showed significantly reduced TF production compared to healthy controls, quantified by WL.^16^ The finding of a strong positive correlation between WL and total protein concentration aligns with previous findings.^24, 25^

We further examined the frequency of a 24bp-duplication polymorphism in the CHIT1 gene, which is present in about one third of the European population^23^. Although homozygotes were detected only among ALS patients in this cohort and the polymorphism showed a trend towards a higher frequency in this group, the difference did not reach statistical significance. Previous studies have reported varying findings regarding this association.^13, 26^ The polymorphism is known to result in lower levels of CHIT1 in serum and CSF due to increased intracellular degradation^13, 14^, which is confirmed in our data. Notably, we show CHIT1 expression in TF is affected to the same extent by the respective genotype, suggesting that the impact of the polymorphism extends across different biofluids. Because of the largely even distribution of the polymorphism in our cohort, we chose neither to correct for the polymorphism nor to exclude homozygous carriers, an approach chosen by most other studies as well, although the genotype was only rarely analyzed.

Chitinases are established markers of neuroinflammation and known to be increased in the CSF but not the serum of ALS patients^9, 27, 28^, which was confirmed in this study. Importantly, this study, to the best of our knowledge, provides the first insight into the abundances of CHIT1 and CHI3L1 in TF. We show significantly elevated concentrations of CHI3L1 in the TF of ALS patients. CHIT1 levels exhibit the same trend, albeit missing significance. There was no significant correlation of CHIT1 and CH3L1 abundances between TF and CSF. We postulate that these results could become significant with a larger number of participants, particularly given the low number of matching CSF samples available for the present analysis. Interestingly, CHIT1 and CHI3L1 in TF separately, as well as combined, showed a better performance in distinguishing ALS patients from healthy controls using ROC-curve analysis compared to serum. Here, CHI3L1 in TF alone showed the strongest diagnostic performance within TF and even outperformed its counterpart in CSF. Previously, however, CHIT1 was shown to have better discriminating power than CHI3L1 in CSF.^29^ One explanation for the observed inferiority of CHIT1 compared to CHI3L1 in TF in our study might be its low concentration in TF, which resulted in a third of the samples being below the limit of detection.

Considering clinically relevant subgroups, we observed higher CHIT1 values in males in TF only, not in serum or CSF. In the male subgroup, CHIT1 values further correlated with the progression rate of ALS patients. This is in line with studies of CSF data where CHIT1 has been described to be associated with the progression rate.^27, 28^ Additionally, both Chitinases in TF individually and in combination showed a better classifier performance in males compared to both females as well as to the whole cohort. Sex-related differences in ALS have been frequently discussed, but the underlying biological mechanisms are still not completely understood.^4, 30, 31^ Interestingly, males were shown to demonstrate stronger transcriptomic, miRNAomic as well as proteomic deregulations in prefrontal cortex tissue, suggesting a more aggressive disease progression than in females.^4^ Whether the difference observed in this study reflects characteristics of the TF itself or is connected to pathological mechanisms in ALS needs to be further studied.

Lastly, we investigated the additive discriminative power of serum NfL, a well-described biomarker in ALS, in combination with chitinase values of the different biofluids. While the diagnostic accuracy of serum NfL did not increase using the serum abundances of the chitinases, values in CSF, and, importantly, TF, improved the accuracy. This supports the notion that NfL and chitinases reflect different aspects of the disease, namely axonal damage and neuroinflammation, highlighting their complementary potential. Taken together, both serum NfL and TF chitinases may therefore provide a less invasive approach for longitudinal monitoring.

Our study has important limitations. Due to the low concentration of CHIT1 in TF and the resulting limited detectability, a subset of patients’ values could not be measured, and small differences among the patients were potentially missed. In addition, CSF samples collected during the same visit as serum and TF were only available for a limited group of patients.

In conclusion, this study is, to the best of our knowledge, the first to investigate the abundance of chitinases, namely CHIT and CHI3L1, in the TF of ALS patients compared to healthy controls. This supports the potential of TF as a non-invasively collectable biofluid, reflecting similar alterations as in CSF, which are, crucially, not detectable in blood. Further studies using more sensitive analytical techniques and larger, independent cohorts are needed to expand on these findings.

## Declarations

### Author contributions

L.W.: investigation, formal analysis, visualization, methodology, writing - original draft, writing - review & editing. L.-S.S.: methodology, writing - review & editing. N.R.: investigation, writing - review & editing. R.v.H.: investigation, writing - review & editing. V.G.: investigation, writing - review & editing. P.R.: investigation, writing - review & editing. P.J.M.: investigation, writing - review & editing. E.F.: supervision, resources, writing - review & editing. A.F.D.: conceptualization, investigation, formal analysis, methodology, supervision, writing - original draft, writing - review & editing. P.L.: conceptualization, formal analysis, methodology, supervision, resources, writing - original draft, writing - review & editing.

### Funding

P.L. received funding from the DFG (German Research Foundation) under Germany’s Excellence Strategy within the framework of the Munich Cluster for Systems Neurology (EXC 2145 SyNergy – ID 390857198). A.F.D. was supported by the Kommission für Klinische Forschung (KKF) of the TUM School for Medicine and Health (project H-13).

### Competing Interests

The authors declare no competing interests.

### Data availability statement

Original data (including deidentified participant data) is available with the authors upon reasonable request.

### Patient consent for publication

Not applicable.

### Ethics approval

This study involved human participants and was approved by the Ethics review board of the Technical University of Munich (approval numbers: 9/15S, 2021-473-S-KH). Participants gave informed consent to participate in the study before taking part.

## Acknowledgements

The authors thank all patients and caregivers for their participation in the study.

## Legend descriptions

**Table 1: Characteristics of the study population**

Abbreviations: ALS, amyotrophic lateral sclerosis; CHIT1, chitotriosidase; CHI3L1, chitinase-3-like protein 1; TF, tear fluid; CSF, cerebrospinal fluid; NfL, neurofilament light chain; ALS-FRS-R, revised amyotrophic lateral sclerosis functional rating scale; SD, standard deviation.

TF-values were adjusted for total protein concentration.

a: Wilcoxon rank sum test

b: Pearson‘s Chi-squared test

c: Multiple linear regression correcting for age and sex

d: Wilcoxon rank sum exact test

## Notes

### Competing Interest Statement

The authors have declared no competing interest.

