## Supplementary Material for "Chitinases in tear fluid of patients with Amyotrophic Lateral Sclerosis"

**Additional Information for**

**Additional Information - Table of Contents**

Supplementary Methods	3

Supplementary Figures	4

Supplementary Figure S1: Correlation of the total protein concentration [µg/µl] in TF and wetting length [mm/5min]	4

Supplementary Figure S2: CHIT1-values in serum depending on the CHIT1-polymorphism	5

Supplementary Figure S3: Correlation of chitinase levels across all biofluids	6

Supplementary Figure S4: Correlation of chitinase-values in TF with clinical parameters	7

Supplementary Figure S5: Correlation of NfL-values in serum with chitinase-levels in all biofluids	8

Supplementary Figure S6: ROC-analysis of chitinase-values in TF (ALS vs. controls), separated by sex	9

Supplementary Tables	10

Supplementary Table S1: Characteristics of the CSF samples	10

### Supplementary Methods

For TF collection, Schirmer strips were placed in the lower lateral eyelid of each patient’s eye and left in place for 5 min with closed eyes. Afterwards, the strips were removed, put in sample storage tubes, and immediately frozen at -20°C before being transferred to -80°C within one week. For processing, the strips were cut into small parts, soaked with 150 µL of RIPA buffer, and incubated at room temperature for 1 h with constant agitation. The tube containing the strips was then placed into a bigger tube and pierced with a needle to collect the fluid components in the bigger tube. After centrifugation at 16,000 × g for 10 min, the supernatant was transferred into a new tube and filled up with RIPA buffer to reach a final volume of 200 µL.

CSF and serum samples were centrifuged at 1,160 x g for 10 min, aliquoted in polypropylene tubes and stored within 2 h at -80°C.

### Supplementary Figures

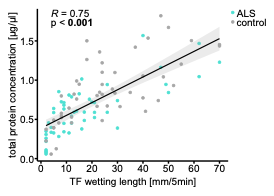

**Supplementary Figure S1: Correlation of the total protein concentration [µg/µl] in TF and wetting length [mm/5min]**

Regression graph for total protein concentration [µg/µl] in TF and wetting length [mm/5min]. The regression line is shown in black with 95% confidence interval as well as all individual data points (turquoise = ALS, *n* = 50; grey = control, *n* = 50). Pearson’s correlation coefficient was calculated and is depicted at the top with the corresponding p-value.

Abbreviations: ALS, amyotrophic lateral sclerosis; TF, tear fluid.

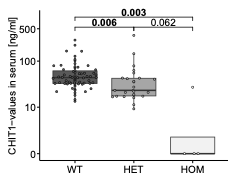

**Supplementary Figure S2: CHIT1-values in serum depending on the CHIT1-polymorphism**

CHIT1-values in serum depending on the CHIT1-polymorphism (wildtype, *n* = 66; heterozygous, *n* = 36; homozygous, *n* = 4). Subgroups were compared by Kruskal-Wallis test followed by Dunn’s post-hoc test adjusted for multiple comparisons with Holm correction. Absolute p-values are depicted. Boxplots show median and 1st and 3rd quartiles as well as all individual data points.

Abbreviations: CHIT1, chitotriosidase; WT, wildtype; HET, heterozygous; HOM, homozygous.

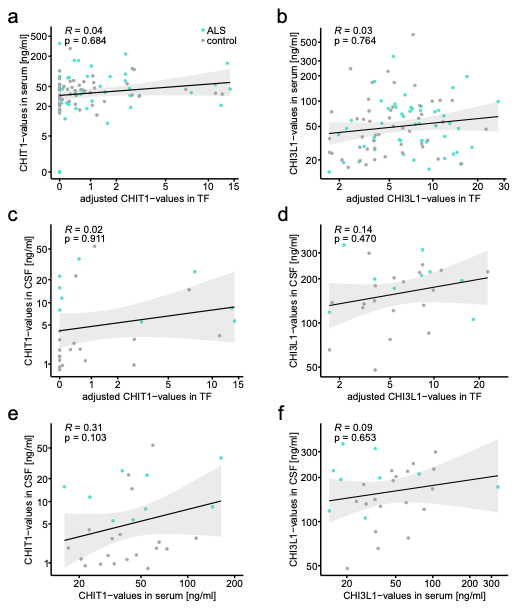

**Supplementary Figure S3: Correlation of chitinase levels across all biofluids**

Regression graphs for CHIT1- and CHI3L1-values between TF and serum (**a**, **b**; *n* = 99), TF and CSF (**c**, **d**; *n* = 28), and serum and CSF (**e**, **f**; *n* = 28), respectively.

For all graphs the regression line is shown in black with 95% confidence interval as well as all individual data points (turquoise = ALS, grey = control). Pearson’s correlation coefficient was calculated and is depicted at the top with corresponding p-values. TF-values were adjusted for total protein concentration.

Abbreviations: CHIT1, chitotriosidase; CHI3L1, chitinase-3-like protein 1; ALS, amyotrophic lateral sclerosis; TF, tear fluid; CSF, cerebrospinal fluid.

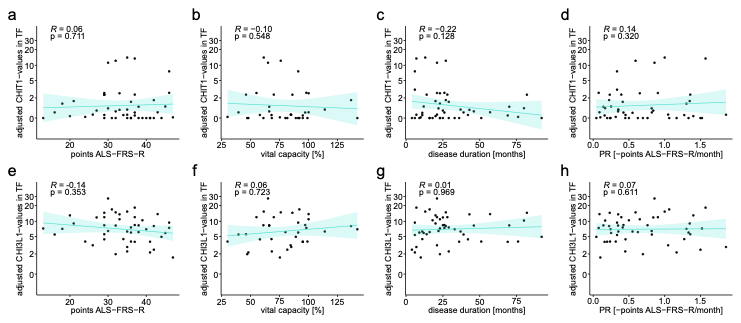

**Supplementary Figure S4: Correlation of chitinase-values in TF with clinical parameters**

**a, b, c, d** Regression graphs for CHIT1-values in TF and ALSFRS-R (**a**; *n* = 47), vital capacity (**b**; *n* = 36), disease duration (**c**; *n* = 50) and progression rate (**d**; *n* = 50).

**e, f, g, h** Regression graphs for CHI3L1-values in TF and ALSFRS-R (**d**; *n* = 47), vital capacity (**e**; *n* = 36), disease duration (**f**; *n* = 50) and progression rate (**h**; *n* = 50).

For all graphs, the regression line is shown in turquoise with a 95% confidence interval as well as all individual data points. Pearson’s correlation coefficient was calculated and is depicted at the top with corresponding p-values. TF-values were adjusted for total protein concentration.

Abbreviations: CHIT1, chitotriosidase; CHI3L1, chitinase-3-like protein 1; ALS, amyotrophic lateral sclerosis; TF, tear fluid; ALS-FRS-R, revised amyotrophic lateral sclerosis functional rating scale; PR, progression rate.

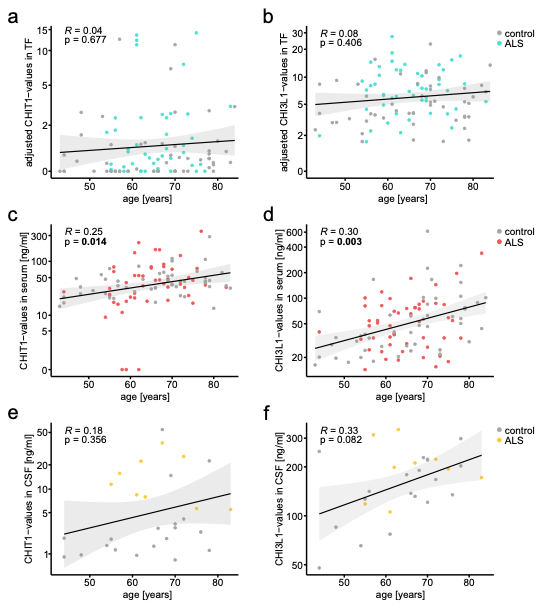

**Supplementary Figure S5: Correlation of NfL-values in serum with chitinase-levels in all biofluids**

Regression graphs for NfL-values in serum and CHIT1-values as well as CHI3L1-values in TF (**a,** **b**; *n* = 96), in serum (**c, d**; *n* = 96) and in CSF (**e, f**; *n* = 25)

For all graphs, the regression line is shown in black with a 95% confidence interval as well as all individual data points (turquoise, red, yellow = ALS, grey = control). Pearson’s correlation coefficient is depicted at the top with corresponding p-values. TF-values were adjusted for total protein concentration.

Abbreviations: CHIT1, chitotriosidase; CHI3L1, chitinase-3-like protein 1; NfL, neurofilament light chain; ALS, amyotrophic lateral sclerosis; TF, tear fluid; CSF, cerebrospinal fluid.

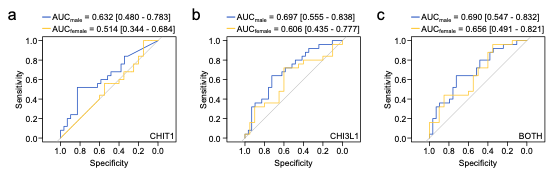

**Supplementary Figure S6: ROC-analysis of chitinase-values in TF (ALS vs. controls), separated by sex**

**a, b, c** ROC-analysis of CHIT1 (**a**), CHI3L1 (**b**) and both chitinases (**c**), divided in the male (blue line; *n* = 54) and female (yellow line; *n* = 45) subgroups. AUC values with the 95% confidence interval are depicted.

Abbreviations: CHIT1, chitotriosidase; CHI3L1, chitinase-3-like protein 1; ALS, amyotrophic lateral sclerosis; TF, tear fluid; ROC, receiver operating characteristic; AUC, area under the curve.

### Supplementary Tables

|  |  | **control** | **ALS** | **p** |
| --- | --- | --- | --- | --- |
| N |  | 19 | 9 |  |
| cell count / µl | mean ± SD | 1.79 ± 1.36 | 1.89 ± 3.10 | 0.271^A^ |
|  | median  (min - max) | 2.0  (0 - 5) | 1.0  (0 - 10) |  |
| glucose [mg/dl] | mean ± SD | 74.79 ± 11.53 | 78.33 ± 16.33 | 0.622^A^ |
|  | median  (min - max) | 71.0  (60 - 102) | 75.0  (64 - 117) |  |
| lactate [mmol/l] | mean ± SD | 1.88 ± 0.313 | 1.78 ± 0.30 | 0.430^B^ |
|  | median  (min - max) | 1.89  (1.40 - 2.50) | 1.78  (1.32 - 2.31) |  |
| albumin [CSF/serum ratio] | mean ± SD | 7.77 ± 4.12 | 7.67 ± 4.25 | 0.824^A^ |
|  | median  (min - max) | 7.0  (3.6 - 21.6) | 6.3  (3.8 - 15.6) |  |

##### **Supplementary Table S1: Characteristics of the CSF samples**

Abbreviations: ALS, amyotrophic lateral sclerosis; CSF, cerebrospinal fluid; SD, standard deviation.

A: Wilcoxon rank sum test

B: T-test
